# Use of human albumin solution in a secondary hospital: an observational, retrospective, cross-sectional study

**DOI:** 10.1101/2022.08.23.22279130

**Authors:** Rosa Rodriguez Mauriz, Patricia Monzó, Nuria Rudi Sola, Ares Villagrasa, Laura Borràs Trias

**Author notes:** **Correspondence Address:** Laura Borràs Trias, Pharmacy Department, Hospital General de Granollers, Avda. Francesc Ribas s/n, 08402, Granollers, Barcelona, Spain.

## Abstract

Human albumin solution (HAS) is a plasma-derived product used for a wide variety of clinical indications, although only some of these are supported by solid scientific evidence.

**Objectives:** The primary aim of this study was to analyze the level of evidence supporting the clinical indications for which HAS is used at a secondary care hospital. Secondary objectives were to evaluate dosing appropriateness and perform a subanalysis of the internal medicine department.

**Methods:** Retrospective, observational, multidisciplinary study of adults who received at least one dose of HAS during 2019. We analyzed sociodemographic, clinical, laboratory, and pharmacotherapy variables. The indications for which HAS was used were classified into the four evidence-based categories established by the American Society for Apheresis: high priority (category I), reasonable evidence (II), weak evidence (III), and treatment not recommended (IV). Clinical guideline recommendations were used to evaluate dosing appropriateness.

**Results:** The study population comprised 142 patients (41% women), mean (SD) age of 66 (14) years. The main admission diagnoses were decompensated cirrhosis (32%) and septic shock (31%). In total, 223 courses of HAS were prescribed. The main indications were treatment of anasarca and hypoalbuminemia (32%), prevention of paracentesis-induced circulatory dysfunction after large-volume paracentesis (17%), resuscitation in septic shock (13%), and treatment of protein malnutrition (9%). Just 26% of hospital-wide indications were supported by strong evidence (category I); 13% category II, 8% category III, and 53% category IV. In the internal medicine department, 36% of indications were in category I, 16% in category III, 48% in category IV. An appropriate dosing regimen was used in just four category I indications.

**Conclusions:** A large proportion of HAS indications at our hospital are supported by weak evidence. Training to promote the rational use of HAS is needed. Protocols and local clinical guidelines could help standardize and optimize its use.

## INTRODUCTION

Human serum albumin accounts for approximately 50% (3.5-5 g/dL) of the total protein in human plasma (1,2). It is the main modulator of fluid distribution between body compartments and is responsible for approximately 75% of plasma oncotic pressure (3). Sixty percent of its oncotic properties are due to direct osmotic effects related to the molecular mass of the protein, while 40% are due to water retention, as the molecule carries a negative charge at physiological pH and therefore attracts sodium ions and other active cations (the Gibbs-Donnan effect) (4).

Albumin also has non-oncotic properties, most of which are related to its molecular structure and conformation. These secondary functions include antioxidant activity, endothelial stabilisation, solubilisation, antithrombotic effects, anti-inflammatory effects, capillary permeability, and binding and transport of numerous plasma constituents and drugs (1–5).

Human albumin solution (HAS) is a plasma-derived product and one of the main limiting factors to its therapeutic use is the limited supply of donors. The production process is also complex, as albumin needs to be extracted from blood, purified, and then formulated into the final product. Moreover, because of the potential risk of viral transmission, minimum EU safety and quality standards must be met, including screening for markers of infection among donors and steps to inactivate or remove any viruses during the production process. Scarcity of donors and manufacturing complexity have made HAS a costly product with limited availability (6).

HAS is currently used for a wide variety of clinical indications, although only some of these are supported by solid scientific evidence. Others are more controversial or scientific evidence has shown that they are inappropriate (4).

In a randomised trial comparing HAS and isotonic saline in cirrhotic patients with acute (grade II-IV of West-Haven criteria) hepatic encephalopathy, HAS was associated with significantly improved survival at 90 days, although no differences were observed for resolution of encephalopathy (7). In a multicentre, randomised, parallel, open-label trial comparing standard care with standard care plus HAS in outpatients with ascites at 33 Italian hospitals, weekly HAS infusions were associated with a significant reduction in the incidence of infections and kidney dysfunction and a lower probability of death (8). Conflicting results, however, have been reported. In a randomised trial involving 777 patients with decompensated cirrhosis and an albumin level <30 g/L, HAS infusion produced no significant benefits over standard care in terms of infection rates, kidney dysfunction, or mortality (9).

International clinical practice guidelines recommend using HAS to treat hepatorenal syndrome (HRS) and prevent paracentesis-induced circulatory dysfunction (PICD) and renal failure induced by spontaneous bacterial peritonitis (SBP) (10,11). The strongest evidence for the use of HAS is for fluid resuscitation in patients with septic shock when crystalloids or non-protein colloids are ineffective or contraindicated (12) and for the treatment or prevention of severe clinical complications in patients with advanced cirrhosis (3,11,13). Robust evidence, however, is lacking for other clinical indications. Rising manufacturing costs and decreasing volumes have increased the cost of HAS and led to supply shortages. Demand, however, has not fallen and the product is still widely used. Clinical and economic evaluations are necessary to ensure a more rational use of HAS in clinical practice (2,3).

HAS usage is high at our hospital, a secondary care centre in Barcelona, Spain. Our hypothesis was that this product was being overused or misused. We designed a study to examine real-life HAS use at our hospital and analyze the level of evidence supporting its prescription in different cases. Secondary aims were to determine the appropriateness of the dosing regimens used for indications with the strongest evidence and perform a subanalysis of HAS use in the internal medicine department.

## METHODS

### Study population

We conducted a single-centre, observational, retrospective, cross-sectional study of all patients aged >18 years who received at least one dose of HAS during hospitalisation or specialised outpatient or emergency care at Hospital General de Granollers in Barcelona, Spain, during 2019. The hospital is a secondary care hospital that serves a population of approximately 399 900 people. The study protocol was approved by the hospital’s research ethics committee.

### Variables

We collected the following data: sociodemographic variables (age, sex), clinical variables (admission diagnosis, previous history of SBP or active infection), laboratory data (serum sodium, albumin, and creatinine levels before HAS administration), number of HAS courses prescribed, ordering provider (department), and pharmacotherapy variables (clinical indications and supporting evidence, doses and treatment duration, and appropriateness of dosing regimens used in indications with the strongest level of evidence).

The indications for which HAS was used were classified into the four evidence-based categories established by the American Society for Apheresis: category I (high-priority use), category II (use supported by reasonable evidence but other treatment options available), category III (weak evidence base), and category IV (use not recommended).

To assign the indications to each category, we conducted a literature search in PubMed and reviewed the guidelines of the American Association for the Study of Liver Diseases (10), the European Association for the Study of the Liver (EASL) Clinical Practice Guidelines for the management of patients with decompensated cirrhosis (1) and the summary of product characteristics for HAS (14). The levels of evidence supporting the guideline recommendations were taken into account to assign each indication to a given category. The results are shown in Table 1.

**Table 1.**
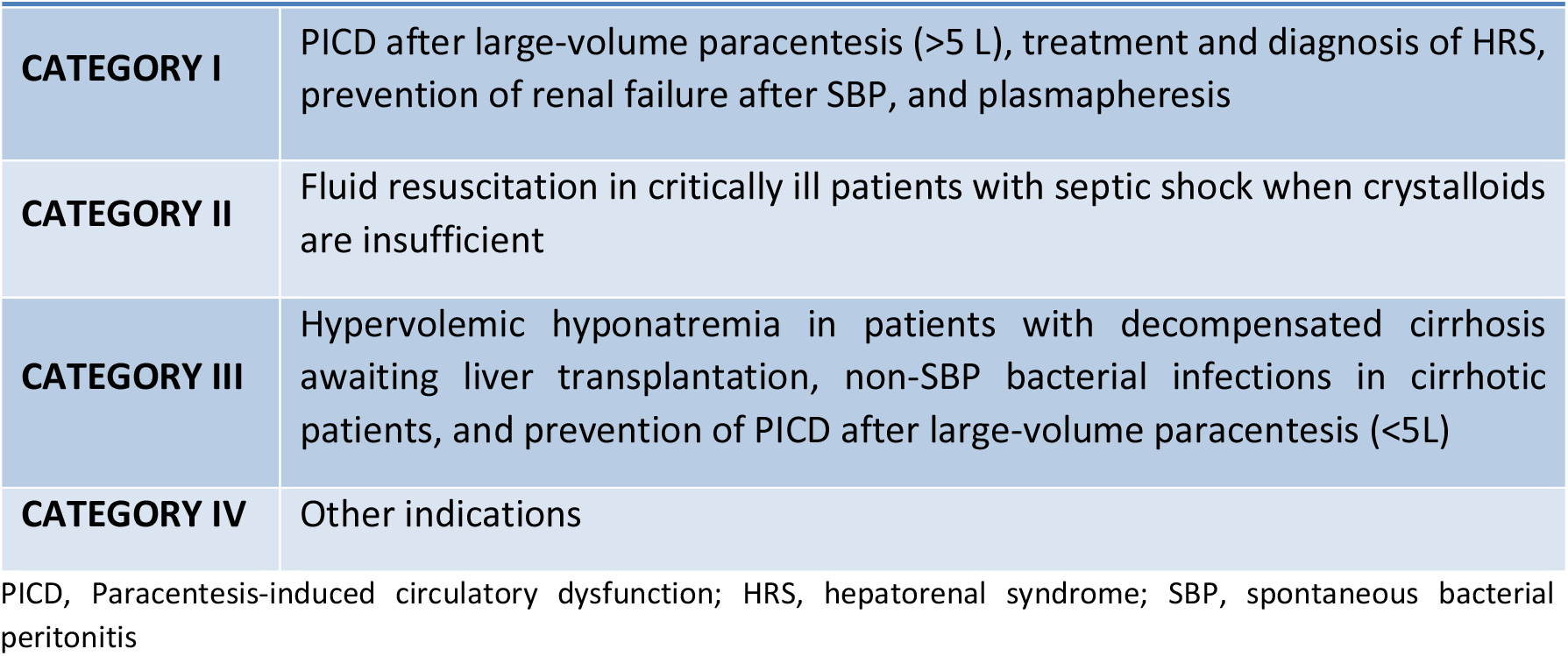
Indications for the use of HAS by evidence-based category

The dosing regimens recommended by the EASL were used as a reference to assess the appropriateness of the regimens prescribed at our hospital. Recommended regimens are 8 g of HAS per litre of ascites removed to prevent PICD after large-volume paracentesis; 1.5 g of HAS per kilogram of body weight at diagnosis followed by 1 g/kg on day 3 to decrease the risk of HRS in SBP; and a fluid challenge consisting of 20%-25% intravenous HAS 1 g/kg/d for 2 days following a diagnosis of acute kidney injury (AKI), together with withdrawal of diuretics and temporary discontinuation of non-selective β blockers. The recommended treatment for AKI-HRS consists of vasoconstrictors plus HAS 20-40 g/d and reversal of precipitating factors. This treatment should be maintained until creatinine levels fall below 1.5 mg/dL or for a maximum of 14 days in cases of partial or no response.

### Statistical analysis

A descriptive statistical analysis was performed. Quantitative variables are expressed as mean (SD) for normally distributed variables and median (IQR) for non-normally distributed variables. Qualitative variables are expressed as percentages.

The analyses were performed in STATA v13.

## RESULTS

We studied 142 patients with a mean (SD) age of 66 (14) years; 41% were women (*n* = 58).

The main admission diagnoses were decompensated cirrhosis (32%; *n*=45), septic shock (31%; *n*=44), haemorrhagic shock (5%; *n*=7), and respiratory infection (4%; *n*=5). Just 5% of patients (*n*=7) had a previous SBP, while 48% (*n*=68) had a previous active infection. Mean serum levels before HAS infusion were 2.5 (0.6) mg/dL for albumin, 1.44 (1.09) mg/dL for creatinine, and 138 mEq/L for sodium (6).

In total, 223 courses of HAS were prescribed; the largest ordering provider was the internal medicine department (38%; *n*=85), followed by intensive care (31%; *n*=69), the internal medicine outpatient clinic (9%; *n*=21), and general surgery (8%; *n*=18).

The demographic and clinical characteristics of the patients are summarised in Table 2.

**Table 2.** Characteristics of included patients

| Table 2 Characteristics of included patients |  | N | % |
| --- | --- | --- | --- |
| Mean (SD) age, y |  | 66 (14) |  |
| Sex | Male | 84 | 59 |
|  | Female | 58 | 41 |
| Mean (SD) serum albumin level before HAS infusion, mg/dL* |  | 2.5 (0.6) |  |
| Mean (SD) serum creatinine level before HAS infusion, mg/dL* |  | 1.44 (1.09) |  |
| Mean serum sodium level before HAS infusion, mEq/L* |  | 138 (6) |  |
| Main admission diagnoses* | Decompensated cirrhosis | 45 | 32 |
|  | Septic shock | 44 | 31 |
|  | Haemorrhagic shock | 7 | 5 |
|  | Respiratory infection | 5 | 4 |
| Previous SBP* |  | 7 | 5 |
| Previous active infection* |  | 68 | 48 |
| Main ordering provider* | Internal medicine | 85 | 38 |
|  | Intensive care | 69 | 31 |
|  | Internal medicine outpatient clinic | 21 | 9 |
|  | General surgery | 18 | 8 |

\*Values are expressed per course of HAS prescribed (n = 223)
SD, standard deviation; HAS, Human albumin solution; SBP, Spontaneous bacterial peritonitis.

### Primary outcomes

#### Indications and grade of evidence

The main indications for hospital-wide HA use were treatment of anasarca with hypoalbuminemia (32%; *n*=71), prevention of PICD after large-volume paracentesis (17%; *n*=38), fluid resuscitation in critically ill patients with septic shock (13%, *n*=29), and treatment of protein malnutrition (9%; *n*=20) (Table 3).

**Table 3.** Indications

| Table 3 Indications | N | % |
| --- | --- | --- |
| Treatment of anasarca with albumin < 3.5g/dL | 71 | 32 |
| Prevention of PICD after paracentesis > 5 L | 38 | 17 |
| Fluid resuscitation in critically ill patients | 28 | 13 |
| Treatment of protein malnutrition | 19 | 9 |
| Prevention of PICD after paracentesis < 5 L | 14 | 6 |
| Treatment and diagnosis of HRS | 12 | 5 |
| Treatment of HE in cirrhotic patients | 9 | 4 |
| Management of ascites | 8 | 4 |
| Prevention of renal failure after SBP | 7 | 3 |
| Others | 17 | 7 |

PICD, paracentesis-induced circulatory dysfunction; HRS, hepatorenal syndrome; HE, hepatic encephalopathy; SBP, spontaneous bacterial peritonitis

Overall, there were 57 category I indications (26%), 28 category II indications (13%), 19 category III indications (8%), and 119 category IV indications (53%).

### Secondary outcomes

#### Dosing regimen

The dosing regimen used in category I indications was considered appropriate in just 7% of cases (*n*=4). Treatment duration was appropriate in 63% of cases (*n*=35) but just 14% of the doses used (*n*=8) were appropriate.

#### Internal medicine subanalysis

Fifty-seven patients on the internal medicine ward received 85 courses of HAS during 69 admissions. The main diagnoses on admission were decompensated cirrhosis (42%; *n*=29), septic shock (12%; *n*=8), and respiratory infection (7%; *n*=5). The baseline characteristics of this subgroup of patients are summarised in Table 4.

**Table 4.** Characteristics of internal medicine patients

| Table 4 Characteristics of internal medicine patients |  | N | % |
| --- | --- | --- | --- |
| Sex | Male | 37 | 65 |
|  | Female | 20 | 35 |
| Age |  | 64 (13) |  |
| Mean (SD) serum albumin level before HAS infusion, mg/dL* |  | 2.62 (0.56) |  |
| Mean (SD) serum creatinine level before HAS infusion, mg/dL* |  | 1.44 (1.10) |  |
| Mean serum sodium level before HAS infusion, mEq/L* |  | 137.6 (5.90) |  |
| Main admission diagnoses | Decompensated cirrhosis | 29 | 42 |
|  | Septic shock | 8 | 12 |
|  | Respiratory infection | 5 | 7 |
|  | Heart failure | 4 | 6 |
|  | Haemorrhagic shock | 3 | 4 |
|  | Infectious diarrhoea | 3 | 4 |
|  | AKI | 3 | 4 |
|  | Alcoholic hepatitis | 3 | 4 |
| Previous active infection | Yes | 21 | 25 |
|  | No | 61 | 72 |

SD, standard deviation; HAS, Human albumin solution; AKI, Acute kidney injury.

The main indications for the use of HAS in hospitalised internal medicine patients were treatment of anasarca with hypoalbuminemia (25%; *n*=21), prevention of PICD after large-volume paracentesis (19%; *n*=16), and treatment of hepatic encephalopathy in cirrhotic patients (9%; *n*=8) (Table 5).

**Table 5.** Indications for the use of HAS in hospitalised internal medicine patients

| Table 5 Indications for the use of HAS in hospitalised internal medicine patients | N | % |
| --- | --- | --- |
| Treatment of anasarca with albumin < 3.5g/dL | 21 | 25% |
| Prevention of PICD after paracentesis > 5 L | 16 | 19% |
| Treatment of protein malnutrition | 8 | 9% |
| Treatment of HE in cirrhotic patients | 7 | 8% |
| Treatment and diagnosis of HRS | 7 | 8% |
| Management of ascites | 6 | 7% |
| Prevention of renal failure after SBP | 6 | 7% |
| Prevention of PICD after paracentesis < 5 L | 5 | 6% |
| Others | 9 | 11% |

HAS, Human albumin solution; PICD, Paracentesis-induced circulatory dysfunction; SBP, spontaneous bacterial peritonitis; HE, Hepatic encephalopathy; HRS, Hepatorenal syndrome; SBP, Spontaneous bacterial peritonitis.

Thirty-six percent (*n*=29) of the indications fell into category I, 16% (*n*=9) into category II, and 48% (*n*=47) into category IV.

Treatment duration was appropriate in 39% of cases (*n*=11), while dose was appropriate in just 18% (*n*=5). Overall, 7% (*n*=2) of the dosing regimens used in the internal medicine department were appropriate.

## DISCUSSION

HAS is used for a wide variety of indications at our hospital, but not all these are supported by solid scientific evidence. Just 26% of the indications corresponded to high-priority use, and over 50% can be considered inappropriate (category IV). Routine use of HAS to treat anasarca with hypoalbuminemia, for example, is not supported by the evidence, yet this indication accounted for almost one-third of HAS prescriptions in our series. In addition, just 7% of the dosing regimens used for category I indications were congruent with guideline recommendations.

*Foroughinia et al* (15) reported an inappropriate HAS use rate of 87.3% at a hospital in Shiraz, Iran, where the main reason for HAS misuse was nephrotic syndrome without hypoalbuminemia. On analyzing HAS prescribing practices at a hospital in Istanbul, Turkey, *Üyetürk et al* (16) found that 50.4% of prescriptions were inappropriate. In this case, hypoalbuminemia and nutritional support were the main reasons for misuse (33.6 %). In another Iranian study, *Talasaz et al*. (17) found that 36.2% of HAS prescriptions at a hospital in Tehran were inappropriate. Findings are difficult to compare across studies due to different definitions of appropriate use. *Vargas et al* (18), for example, considered that HAS should not be used to prevent HRS, while *Foroughinia* et al considered that it should not be used to treat cirrhosis in patients with refractory ascites unless they have a serum albumin level <2 g/L.

Our findings for HAS use in the internal medicine department were similar to our overall results, with just 36% of indications supported by strong evidence. The other indications had a weak evidence base (16%) or were not recommended (48%). These figures are surprising considering that the three category I indications (PICD after large-volume paracentesis, treatment and diagnosis of HRS, and prevention of renal failure after SBP) are common internal medicine conditions. Nonetheless, because almost 50% of HAS prescriptions at our hospital were ordered by the internal medicine department, we did not expect to see significant differences between hospital-wide and internal medicine prescribing practices.

HAS is a blood product derived from human plasma through fractionation and purification processes. It is therefore a costly product with a limited supply, and proper management is crucial (6,19). The findings of this study highlight the need to train prescribers in the rational use of albumin and closely monitor its use. They also reveal the need for proper management of malnutrition and hypoalbuminemia, as these were predominant conditions in our population (mean ± SD albumin 2.5 (0.6) mg/dl) and one of the main reasons for HAS prescription.

The main limitations of our study are those inherent to a single-centre, observational, retrospective design and the lack of solid scientific evidence and agreement on the use of HAS in different indications. We only analyzed the use of HAS from a clinical perspective, but in future studies it would be interesting to assess the economic impact of inappropriate use.

In conclusion, a considerable number of HAS uses at our hospital are supported by little or no evidence. More randomised controlled trials are needed to provide evidence on HAS use for different indications, as robust data are lacking. There is also a need for training to promote rational prescribing practices and for local clinical guidelines and protocols to standardize and optimize its use.

## Data Availability

All data produced in the present study are available upon reasonable request to the authors

## Acknowledgements

The authors thank Elisabet Deig Comerma and Rebeca Acal Arias for their support in this work.

## Contributors

LBT, AV, RRM made substancial contributions to the concept and design of the manuscript. LBT, RRM, PM, AV made substancial contributions to acquisition, analysis and interpratation of data. RRM, PM drafted the manuscript. LBT, AV made critical revisions for important intellectual content and supervised the manuscript. NRS is the guarantor. All authors read, discussed and approved the final manuscript.

## Ethics approval

The study was approved by the Ethics Committee of the Hospital General de Granollers, Spain.

## Patient consent

Not required.

## REFERENCES

1. Walayat S, Martin D, Patel J, Ahmed U, N. Asghar M, Pai AU, et al. Role of albumin in cirrhosis: from a hospitalist’s perspective. J Community Hosp Intern Med Perspect. 2017;7(1):8–14.

2. Caraceni P, Domenicali M, Tovoli A, Napoli L, Ricci CS, Tufoni M, et al. Clinical indications for the albumin use: Still a controversial issue. Eur J Intern Med. 2013;24(8):721–8.

3. Caraceni P, Tufoni M, Bonavita ME. Clinical use of albumin. Blood Transfus. 2013;11(SUPPL. 4).

4. Rozga J, Piatek T, Małkowski P. Human albumin: Old, new, and emerging applications. Ann Transplant. 2013;18(1):205–17.

5. Evans T. Review article : albumin as a drug — biological effects of albumin unrelated to oncotic pressure. Aliment Pharmacol Ther. 2002;16(Suppl. 5):6–11.

6. Committee for medicinal products for human use (CHMP)-European Medicines Agency. Guideline on plasma-derived medicinal products [Internet]. 2011. Available from: https://www.ema.europa.eu/en/documents/scientific-guideline/guideline-plasma-derived-medicinal-products_en.pdf

7. Simón-Talero M, García-Martínez R, Torrens M, Augustin S, Gómez S, Pereira G, et al. Effects of intravenous albumin in patients with cirrhosis and episodic hepatic encephalopathy: A randomized double-blind study. J Hepatol. 2013;59(6):1184–92.

8. Caraceni P, Riggio O, Angeli P, Alessandria C, Neri S, Foschi FG, et al. Long-term albumin administration in decompensated cirrhosis (ANSWER): an open-label randomised trial. Lancet. 2018;391(10138):2417–29.

9. China L, Freemantle N, Forrest E, Kallis Y, Ryder SD, Wright G, et al. A Randomized Trial of Albumin Infusions in Hospitalized Patients with Cirrhosis. N Engl J Med. 2021;384(9):808–17.

10. American Association for the Study of Liver Diseases (AASLD). Practice Guideline: Management of Adult Patients with Ascites Due to Cirrhosis. 2012.

11. Angeli P, Bernardi M, Villanueva C, Francoz C, Mookerjee RP, Trebicka J, et al. EASL Clinical Practice Guidelines for the management of patients with decompensated cirrhosis. J Hepatol. 2018;69(2):406–60.

12. Rhodes A, Evans LE, Alhazzani W, Levy MM, Antonelli M, Ferrer R, et al. Surviving Sepsis Campaign: International Guidelines for Management of Sepsis and Septic Shock: 2016. Vol. 43, Intensive Care Medicine. Springer Berlin Heidelberg; 2017. 304–377 p.

13. Garcia-Martinez R, Caraceni P, Bernardi M, Gines P, Arroyo V, Jalan R. Albumin: Pathophysiologic basis of its role in the treatment of cirrhosis and its complications. Hepatology. 2013;58(5):1836–46.

14. European Medicines Agency. Summary of Product Characteristics (SmPC) for human albumin solution [Internet]. 2019. Available from: https://www.ema.europa.eu/en/documents/scientific-guideline/guideline-core-summary-product-characteristics-human-albumin-solution-revision-3_en.pdf

15. Foroughinia F, Mazraie S. Investigating the use of human albumin in a non-teaching hospital in Iran. Iran J Pharm Res. 2017;16(2):814–9.

16. Üyetürk U, Özbey G, Üyetürk U, Usul Afşar CYS. Evaluation of the use of human albumin solution in Gaziosmanpasa University faculty of medicine hospital. Yeni Tıp Derg. 2010;27:165–8.

17. Talasaz AH, Jahangard-Rafsanjani Z, Ziaie S, Fahimi F. Evaluation of the pattern of human albumin utilization at a University affiliated hospital. Arch Iran Med. 2012;15(2):85–7.

18. Vargas E, De Miguel V, Portolés A, Avendaño C, Ambit MI, Torralba A, et al. Use of albumin in two Spanish university hospitals. Eur J Clin Pharmacol. 1997;52(6):465–70.

19. Yazdani MS, Retter A, Maggs T, Li P, Robson MG, Reid C, et al. Where does the Albumin go? Human Albumin Solution usage following the implementation of a demand management programme. Transfus Med. 2017 Jun 1;27(3):192–9.

